# Computed tomography-derived normative values and z-scores of right ventricular outflow tract structures in the pediatric population

**DOI:** 10.1101/2023.07.31.23293466

**Authors:** Natalie Soszyn, Justin Schweigert, Salvador R. Franco, Gareth J. Morgan, Max Mitchell, Jenny E. Zablah

## Abstract

**Background:** Recent advances in available percutaneous device technology require accurate measurements and quantification of relationships between right ventricular outflow tract (RVOT) structures in children with and without congenital heart disease to determine device suitability. To date, no population study has described normal reference ranges of these measurements by computed tomography (CT). We aimed to establish normative values for four CT-derived measurements between RVOT structures from a heterogeneous population without heart disease and develop z-scores useful for clinical practice.

**Methods:** Patients without heart disease who underwent cardiac CT between April 2014 and February 2021 at Children’s Hospital Colorado were included. Distance between the right ventricular (RV) apex to pulmonary valve (PV), PV to pulmonary trunk bifurcation, and bifurcation to the right and left pulmonary artery were measured. Previously validated models were used to normalize the measurements and calculate Z-scores. Each measurement was plotted against BSA and Z-scores distributions were used as reference lines.

**Results:** Three-hundred sixty-four healthy patients with a mean age of 8.8 years (range 1–21), 58% male, and BSA of 1 m^2^ (range 0.4–2.1) were analyzed. The Haycock formula was used to present data as predicted values for a given BSA and within equations relating each measurement to BSA. Predicted values and Z-score boundaries for all measurements are presented.

**Conclusion:** We report CT-derived normative data for four measurements between RVOT structures from a heterogenous cohort of healthy children. Knowledge of this normative data will be useful in both determining device fit and customising future devices to accommodate the diverse pediatric size range.

**Clinical Perspective:** Recent advances in advanced percutaneous technology require accurate measurements and quantification of relationships between RVOT structures to determine patient selection and device suitability. The availability of a robust range of normal reference values can provide valuable information for current device selection and future development of customized devices for the RVOT tailored to accommodate the broad size, anatomic and physiologic heterogenicity seen in the pediatric population. We report CT-derived normative data for four measurements between cardio-pulmonary structures from a heterogenous cohort of healthy children and young adults. This data may be useful in planning surgical or catheter-guided interventions in the management of children with congenital heart disease.

## Introduction

Accurate measurement of the distance between right ventricular outflow tract (RVOT) structures is vital for management of critically unwell children and young adults with and without congenital heart disease. Recent advances in advanced percutaneous technology require accurate measurements and quantification of relationships between RVOT structures to determine patient selection and device suitability.

Pediatric mechanical circulatory support has advanced over the last decade with momentum building in the development and implantation of durable ventricular assist devices (VADs) in children with increasingly complex anatomy. (1) Deterioration of right ventricular function is an acute event observed in a significant proportion of patients after left ventricular assist device (LVAD) implantation, with approximately 25% of patients requiring biventricular assist device (BiVAD) support and is associated with higher adverse outcomes in the pediatric compared to the adult population. (1, 2) There is a burgeoning interest in developing devices specifically customized for the right ventricle to support a wide range of patient sizes, cardiac anatomy and physiology.

In patients with congenital heart disease, transcatheter pulmonary valve replacement (TPVR) has emerged as a minimally invasive approach to managing RVOT dysfunction due to significant pulmonary valve stenosis or regurgitation. (3–5) Pre-procedural imaging is imperative for identifying patient suitability and device sizing. (6) Similar to trans-catheter aortic valve replacement, cross-sectional CT imaging has been shown to be a useful tool in procedural planning owing to the high isotropic spatial resolution, multiplanar reconstruction capability and acquisition of volumetric data that can allow for 3D reconstruction and printed models useful for procedural guidance. (7, 8) The heterogenicity of patients referred for TPVR means that standardizing RVOT measurements to determine appropriate valve or valve size has presented a challenge. (7) Knowledge of the relationship between the right ventricle, pulmonary valve and branch pulmonary arteries is important in determining whether there is an appropriate landing zone for valve deployment.

The availability of a robust range of normal reference values can provide valuable information for current device selection and future development of customized devices for the RVOT tailored to accommodate the broad size, anatomic and physiologic heterogenicity seen in the pediatric population. To date, no large population study has outlined normal reference ranges of these distance measurements by CT in healthy pediatric patients, only normative measurements of the pulmonary valve annulus are available. We aimed to establish normative values for four CT-derived measurements between RVOT structures from a heterogeneous population without heart disease and develop z-scores useful for clinical practice.

## Methods

### Study population

All patients who underwent a technically adequate cardiac CT completed between April 2014 and February 2021 at the Children’s Hospital Colorado were considered for the study. Patients with evidence of congenital or acquired heart disease were excluded. Demographic information including gender, ethnicity, and age at the time of the CT were collected retrospectively from electronic medical records. The Haycock formula was used in calculation of BSA for all subjects in the study. The study was approved by the Colorado Multi-Institutional Review Board (COMIRB #19-2292).

### CT image acquisition and data analysis

All CT scans were single phase and non-gated. Measurements were retrospectively performed using a dedicated post-processing software (cvi42, Circle Cardiovascular Imaging, Calgary, Alberta, Canada) by a single operator. Measurements were performed at an estimated time of 15-30% of the cardiac cycle. Distances between the apex of right ventricle (RV) to pulmonary valve (PV) (measure 1), PV to bifurcation of pulmonary trunk (measure 2), bifurcation of the pulmonary trunk to the first branch of the right pulmonary artery (RPA) (measure 3), and bifurcation of the pulmonary trunk to the first branch of the left pulmonary artery (LPA) (measure 4) were measured.

Measure 1-3 were obtained using the same slice on the axial view (Figure 1). Firstly, the distance between the RV apex to the pulmonary valve (measure 1) was obtained. A second line was subsequently made perpendicular to the first line that bisected the PV. The distance from the pulmonary valve to the bifurcation of the pulmonary trunk (measure 2) was then obtained by drawing a line perpendicular from the line bisecting the PV to the pulmonary trunk bifurcation. Measure 3 was then obtained by measuring the distance from the PT bifurcation to the to the first branch of the RPA. Finally, using a different axial slice, measure 4 was obtained from the bifurcation of the pulmonary trunk to the first branch of the LPA.

**Figure 1:**
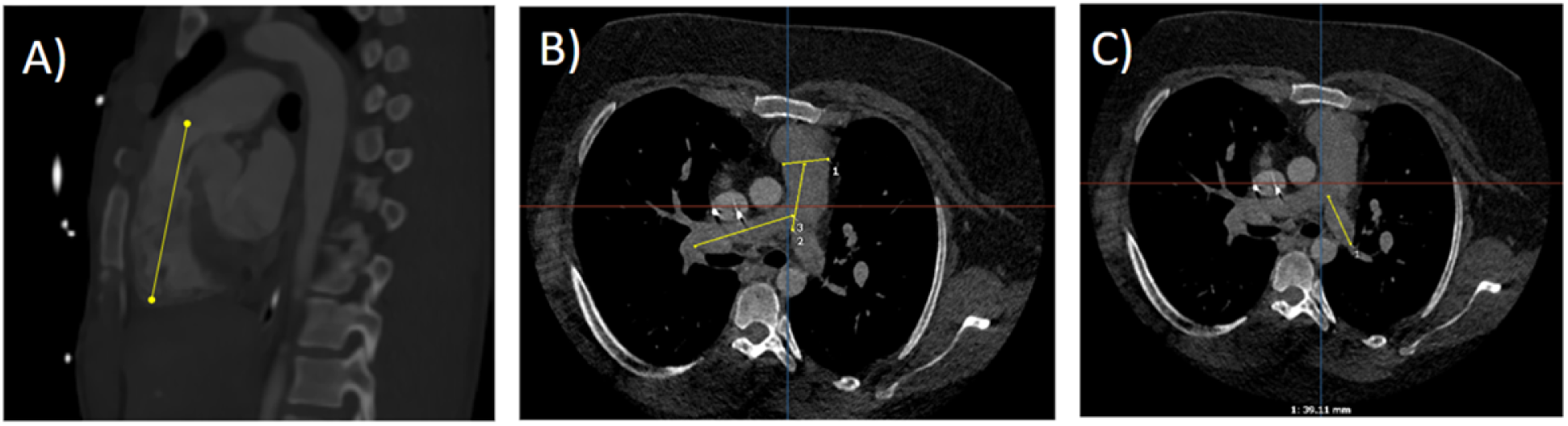
Example of measurements. A. Measure 1 (RV anterior apex to pulmonary valve) obtained in the sagittal plane. B. Measure 2 (PV to PT bifurcation) and measure 3 (PT bifurcation to the first branch of the RPA) obtained in the axial plane by first creating a horizontal line across the plane of the pulmonary valve. C. Measure 4 (PT bifurcation to the first branch of the LPA) in the axial plane.

### Statistical Methods

Statistical analysis was conducted using the statistical software package Stata v.15 (Statacorp LP, College Station, TX). Categorical data was expressed as frequency and percentage. Continuous data was presented as mean (range) if normally distributed and median (interquartile range) if not normally disturbed. A Shapiro-Wilk test was performed to test the null hypothesis that there was no difference between the calculated and standard normal distribution of the continuous variables.

BSA was used as an independent variable in a regression analysis for the predicted mean value of each CT-derived measurement. Each measurement was plotted against BSA and then various non-linear models including logarithmic, exponential, polynomial and quadratic models were evaluated to determine “best fit” with tests for heteroscedasticity applied. A non-linear (polynomial) regression model using each observed measurement as the dependent variable (*x*) and BSA as the predictor (independent variable) was determined as the model of best fit.

Normalized data was then used to calculate *z*-scores. To calculate a *z*-score, we defined the mean for each measurement and the corresponding standard deviation (SD). The *z*-score value conveys the magnitude of deviation from the mean. The *z*-score for each parameter can be calculated as follows:

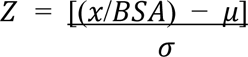

where *x* is the observed measurement, 𝜇 is the mean of the standard population, and 𝜎 is the standard deviation of the sample population.

Standardized *z*-scores are approximately normally disturbed with mean = 0 and SD = 1. Therefore, *z* = 0 corresponds with the estimated mean and *z* = ±1 and ±2 corresponds to ±1 and ±2 SD from the estimated mean. In any normal distribution, 68% of the population is classified within its mean ± 1 SD and 95.4% within its mean ±2 SD.

## Results

### Study population

From a total of 378 who underwent cardiac CT during the defined period, data from 364 healthy children and young adults were included in the study. Fourteen were excluded due to inadequate CT image quality. Patient demographics can be seen in Table 1. The mean age of the study population was 8.8 years ± 6.1 years (median 8 years, IQR 3 – 14 years) and there was 211 males and 153 females. Mean body weight was 32.5 ± 22 kilograms (median 24.3 kg, IQR 14 – 48.1 kg), mean height was 123.5 ± 34 cm (median 121.5 cm, IQR 94 – 155 cm) and mean BSA was 1.1 m^2^ ± 0.5 (median 1.0 m^2^, IQR 0.6 – 1.5). Two-hundred and five patients (56%) identified as being Caucasian, 92 (25%) Hispanic/Latino, 16 (5%) Black/African American, 9 (2%) Asian, 7 (2%) American Indian/Alaska Native, and 1 (2%) Native Hawaiian/Other Pacific Islander. Eighteen patients (4%) identified as being more than one race and 16 (4%) did not specify any ethnicity.

**Table 1:** Patient Demographics.

|  | **Total**<br>(n = 364) |
| --- | --- |
| **Male: Female** | 211 (58%) : 153 (42%) |
| **Age (years)\*** | 8 (3 – 14) |
| **Weight (kg)\*** | 24.3 (14 – 48.1) |
| **Height (cm)\*** | 121.5 (94 – 155) |
| **BSA (m<sup>2</sup>)\*** | 1.0 (0.6 – 1.5) |
| **Ethnicity/Race** |  |
| Caucasian | 205 (56%) |
| Hispanic/Latino | 92 (25%) |
| More than one Race | 18 (5%) |
| Not specified | 16 (4%) |
| Black/African American | 16 (4%) |
| Asian | 9 (2%) |
| American Indian/Alaska Native | 7 (2%) |
| Native Hawaiian/Other Pacific Islander | 1 (2%) |

\*Median (IQR)

### Normal distribution curves

All four measurements were modelled against BSA. A non-linear (polynomial) regression model using BSA provided the best fit for all the data and satisfied the assumption of homoscedasticity and normality of residuals. The measurements plotted against BSA with four curves corresponding to *z*-score = 0, ±1, and ±2 are displayed in Figure 2. The superimposed solid line represents the estimated mean (labelled as *z* = 0). The superimposed dotted lines represent *z* = ±1 and dashed lines *z* = ±2 above and below the mean line.

**Figure 2:**
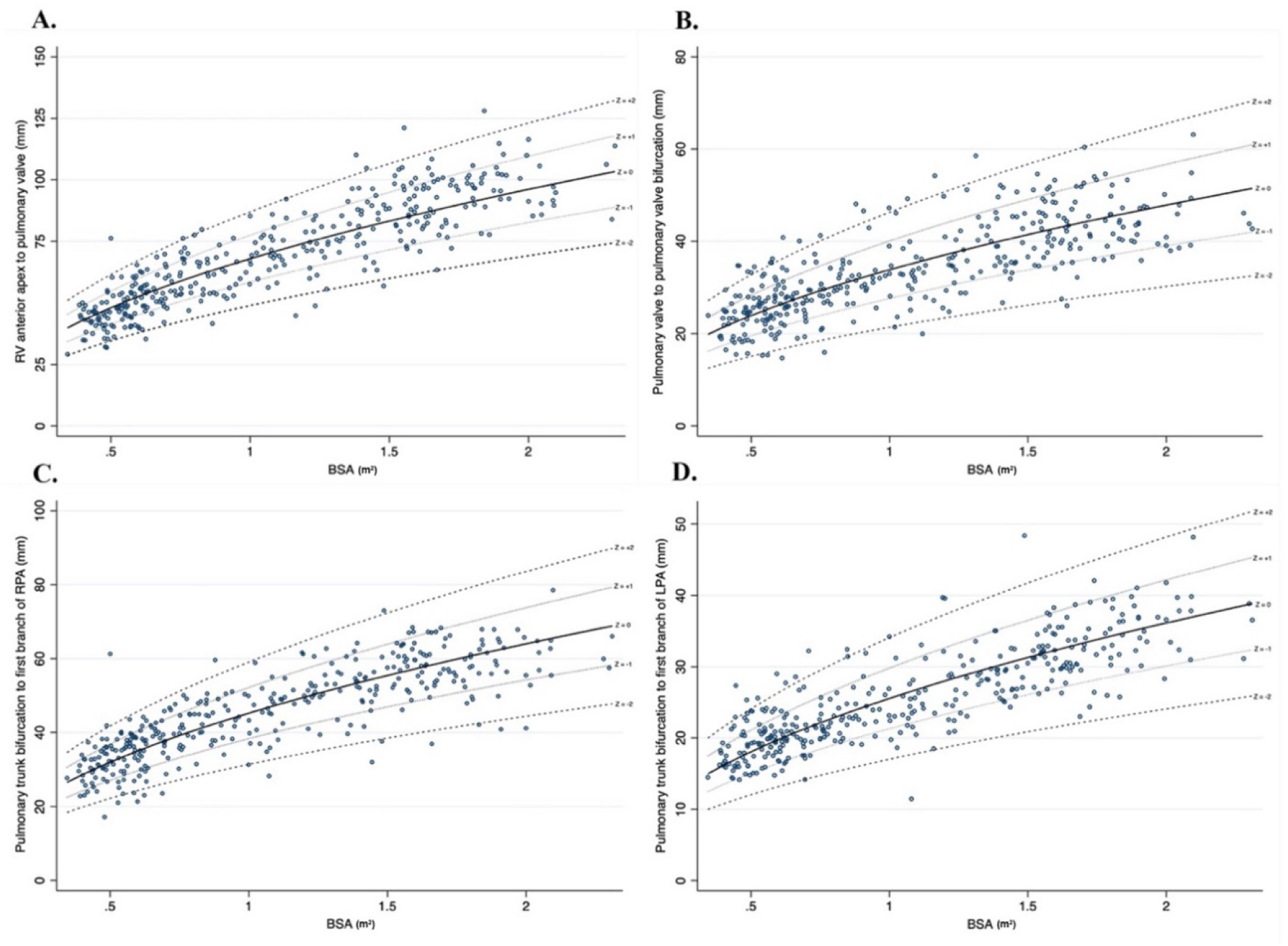
Scatter plot of all four measurements compared to BSA with Z score as reference lines. Panels are separated by measurements as follows: A. RV anterior apex to pulmonary valve; B. pulmonary valve to bifurcation; C. Pulmonary trunk bifurcation to RPA; D. Pulmonary trunk bifurcation to LPA. The solid middle line represents the mean (Z = 0), the two outer dotted lines represent a Z score of +/-1 and two outer dashed lines represent a Z score of +/-2.

### Calculation of Z-scores

Once the best fit relationship was defined, the mean and standard deviation were used at each BSA to determine values for the corresponding *z*-score for each measurement. which is presented in Table 2. The calculated mean and standard deviation for each measurement are provided in Table 3.

**Table 2:** Z-score to BSA for each CT-derived measurement.

| **BSA**<br>**(m<sup>2</sup>)** | **RV anterior apex to**<br>**PV** |  |  | **PV to PT**<br>**bifurcation** |  |  | **PT bifurcation to**<br>**RPA** |  |  | **PT bifurcation to**<br>**LPA** |  |  |
| --- | --- | --- | --- | --- | --- | --- | --- | --- | --- | --- | --- | --- |
|  | **Z score** |  |  | **Z score** |  |  | **Z score** |  |  | **Z score** |  |  |
|  | **-2** | **0** | **+2** | **-2** | **0** | **+2** | **-2** | **0** | **+2** | **-2** | **0** | **+2** |
| 0.5 | 34.6 | 48.1 | 61.5 | 15.1 | 23.9 | 32.8 | 22.2 | 32.0 | 41.8 | 12.0 | 18.1 | 24.1 |
| 0.6 | 37.9 | 52.6 | 67.4 | 16.6 | 26.2 | 35.9 | 24.3 | 35.1 | 45.8 | 13.2 | 19.8 | 26.4 |
| 0.7 | 40.9 | 56.9 | 72.8 | 17.9 | 28.3 | 38.8 | 26.3 | 37.9 | 49.4 | 14.3 | 21.4 | 28.5 |
| 0.8 | 43.8 | 60.8 | 77.8 | 19.1 | 30.3 | 41.4 | 28.1 | 40.5 | 52.9 | 15.2 | 22.9 | 30.5 |
| 0.9 | 46.4 | 64.5 | 82.5 | 20.3 | 32.1 | 43.9 | 29.8 | 42.9 | 56.1 | 16.2 | 24.2 | 32.3 |
| 1.0 | 48.9 | 68.0 | 87.0 | 21.4 | 33.8 | 46.3 | 31.4 | 45.3 | 59.1 | 17.0 | 25.6 | 34.1 |
| 1.1 | 51.3 | 71.3 | 91.2 | 22.4 | 35.5 | 48.6 | 33.0 | 47.5 | 62.0 | 17.9 | 26.8 | 35.7 |
| 1.2 | 53.6 | 74.4 | 95.3 | 23.4 | 37.1 | 50.7 | 34.4 | 49.6 | 64.7 | 18.7 | 28.0 | 28.0 |
| 1.3 | 55.8 | 77.5 | 99.2 | 24.4 | 38.6 | 52.8 | 35.8 | 51.6 | 67.4 | 19.4 | 29.1 | 29.1 |
| 1.4 | 57.9 | 80.4 | 103.0 | 25.3 | 40.0 | 54.8 | 37.2 | 53.5 | 69.9 | 20.2 | 30.2 | 40.3 |
| 1.5 | 59.9 | 83.2 | 106.5 | 26.2 | 41.5 | 56.7 | 38.5 | 55.4 | 72.4 | 20.9 | 31.3 | 41.7 |
| 1.6 | 61.9 | 86.0 | 110.0 | 27.0 | 42.8 | 58.6 | 39.8 | 57.2 | 74.7 | 21.5 | 32.3 | 43.1 |
| 1.7 | 63.8 | 88.6 | 113.4 | 27.9 | 44.1 | 60.4 | 41.0 | 59.0 | 77.0 | 22.2 | 33.3 | 44.4 |
| 1.8 | 65.6 | 91.2 | 116.7 | 28.7 | 45.4 | 62.1 | 42.2 | 60.7 | 79.3 | 22.9 | 34.3 | 45.7 |
| 1.9 | 67.4 | 93.7 | 119.9 | 29.5 | 46.6 | 63.8 | 43.3 | 62.4 | 81.4 | 23.5 | 35.2 | 47.0 |
| 2.0 | 69.2 | 96.1 | 123.0 | 30.2 | 47.9 | 65.5 | 44.4 | 64.0 | 83.6 | 24.1 | 36.1 | 48.2 |

RV = right ventricle; PV = pulmonary valve; PT = pulmonary trunk; RPA = right pulmonary artery; LPA = left pulmonary artery

**Table 3:** Calculated mean and standard deviation for all CT-derived measurements.

| **Measurement** | **Mean ($\mu$)** | **Standard Deviation ($\sigma$)** |
| --- | --- | --- |
| RV anterior apex to pulmonary valve (mm) | 67.96 | 9.52 |
| PV to bifurcation of pulmonary trunk (mm) | 33.84 | 6.23 |
| PT bifurcation to first branch of the RPA (mm) | 45.25 | 6.92 |
| PT bifurcation to first branch of the LPA (mm) | 25.55 | 4.26 |

*Z*-scores using our results can be calculated using two methods. The first is to use the graphs provided to find an approximate corresponding *z-*score for a given observed measurement and BSA. Secondly, Table 3 values can be used to calculate the *z*-score directly and more precisely. A *z-*score calculator is also provided for ease of calculation (Supplementary Material 1). For example, assuming the RV apex to pulmonary valve distance of 48.9 mm and the BSA of a patient is 1.0 m^2^, a z-score can be calculated with the mean of 67.96 mm and defined standard deviation of 9.52 mm as follows:

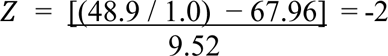

## Discussion

The normal size of right sided cardiac structures including the right ventricle, pulmonary valve annulus, and branch pulmonary artery sizes in children have been well established in the past by echocardiographic, angiographic and even MRI studies. (9–12) Information about the distance between the right ventricle, pulmonary valve, pulmonary artery bifurcation and branch pulmonary arteries in the pediatric population remains scarce. As there is increasing interest in implanting devices in the RVOT, knowledge about the normal range of distance between RVOT structures is important to aid in not only recognizing disease severity but also in patient selection and pre-procedural planning. This is the first study to provide CT-derived, normalized values for four measurements between RVOT structures in a heterogenous population of healthy children and young adults.

The use of BSA as expression of body size in children and the use of regression models to describe the relationship between body growth and the cardiovascular dimensions have been previously validated. (11–13) Similar to other studies, we identified a non-linear relationship between body growth and distances between RVOT structures. In the literature, there is limited description of the relationship between somatic growth and distance between RVOT structures. Our data shows wider variability in all four measurements as BSA increases.

By establishing the ideal relationship between RVOT structures, the results of this study are likely of unique interest to pediatric cardiac surgeons and interventional cardiologists. In presenting normative data, we provide a reference point for physicians in describing disease severity, determining patient selection for procedures, and benchmarking the development of future devices.

In patients with RVOT anomalies, patient selection for TPVR depends on the identification of an appropriate site for implantation within the RVOT that will ensure device stability. Cross-sectional CT imaging has been shown to be a useful tool in procedural planning owing to the high isotropic spatial resolution in all planes and cardiac phases. (7, 16) Shievano et al described five common types of RVOT described by mathematical rules, highlighting the importance of 3D imaging in patient assessment and selection, and that the type of RVOT morphology in combination with RVOT size and distensibility allowed informed selection for currently available balloon expandable valves. (17) The relative distances between the right ventricle, pulmonary valve and branch pulmonary arteries are important in determining whether there is an appropriate landing zone for valve deployment.

There is additionally an increasing interest in using percutaneous VADs in the pediatric population as a less invasive alternative to extracorporeal membrane oxygenation (ECMO). Lasa et al first reported the use of the Impella RP in combination with the Impella CP in a pediatric patient with acute severe biventricular failure. (14) A multi-institutional experience was published in 2021 in adolescents as young as 14 years. (15) Positioning is performed over a guidewire leaving the end of the device (pigtail configuration) at the main pulmonary artery/left pulmonary artery junction, with the exit port in the main pulmonary artery and the inflow port positioned in the inferior vena cava, near the right atrium. (14, 15) The device has a double curved configuration that is ideally suited for the geometry of the right heart in adults and not necessarily for children. For this technology to be applicable to smaller pediatric patients, the device profile and configuration will need to be altered to conform to smaller hearts. Acknowledging this, our normative data provides a complementary tool that will aid in the expanded development of future similar devices for the wide size and anatomic variation seen in the pediatric population.

### Strengths and Limitations

This data are derived from a homogeneous cohort of healthy children considering gender, age, and ethnic background. The study had a retrospective cohort design but had robust statistical methodology. Additionally, measurements were performed by a single operator minimizing interobserver variability and bias. The power of this study may be limited, however, by the fact that only children with a BSA greater than 0.4 m^2^ and less than 2.3 m^2^ were included with a paucity of results greater than 2 m^2^. Even though a regression curve could be extrapolated from our data, we recommend caution for the use of our normative data for children at the extremes of BSA (i.e. less than 0.5 m^2^ and greater than 2 m^2^). Another limitation that requires consideration is the variability of scan techniques and contrast media injection protocols which may influence the accuracy of measurements. CT images also represent an estimated time of 15-30% of the cardiac cycle and could vary between patients. However, only technically adequate CT images were included in analysis minimizing the limitation of multiple varying CT protocols.

## Conclusion

We report CT-derived normative data for four measurements between cardio-pulmonary structures from a heterogenous cohort of healthy children and young adults. This data may be useful in planning surgical or catheter-guided interventions in the management of children with congenital heart disease. Knowledge of this normative data will be useful in customising future devices to accommodate the diverse size range, anatomy, and physiology of the pediatric population.

## Data Availability

The deidentified data underlying this article can be shared on reasonable request to the corresponding author. However, data shall be shared after approval from the Institutional Ethics Committee of Children?s Hospital Colorado.

## Acknowledgements

None

## Sources of Funding

None

## Disclosures

None

## Non-standard Abbreviations and Acronyms

BiVAD: biventricular assist device
BSA: body surface area
CT: computed tomography
LPA: left pulmonary artery
PT: pulmonary trunk
PV: pulmonary valve
RPA: right pulmonary artery
RV: right ventricle
RVOT: right ventricular outflow tract
SD: standard deviation
VAD: ventricular assist device

## Notes

### Competing Interest Statement

The authors have declared no competing interest.

### Funding Statement

No external funding was received.

### Author Declarations

The study was approved by the Colorado Multi-Institutional Review Board (COMIRB #19-2292).

